# Comprehensive Testing Highlights Racial, Ethnic, and Age Disparities in the COVID-19 Outbreak: Epidemiological and Clinical Characteristic of Cases in Utah

**DOI:** 10.1101/2020.05.05.20092031

**Authors:** Sharia M Ahmed, Rashmee U. Shah, Margaret Bale, Jordan B. Peacock, Ben Berger, Alyssa Brown, Sara Mann, William West, Valerie Martin, Valerie Fernandez, Sara Grineski, Benjamin Brintz, Matthew H. Samore, Matthew J. Ferrari, Daniel T. Leung, Lindsay T. Keegan

## Abstract

The United States (US), which is currently the epicenter for the COVID-19 pandemic, is a country whose demographic composition differs from that of other highly-impacted countries. US-based descriptions of SARS-CoV-2 infections have, for the most part, focused on patient populations with severe disease, captured in areas with limited testing capacity. The objective of this study is to compare characteristics of positive and negative SARS-CoV-2 patients, in a population primarily comprised of mild and moderate infections, identified from comprehensive population-level testing. Here, we extracted demographics, comorbidities, and vital signs from 20,088 patients who were tested for SARS-CoV-2 at University of Utah Health clinics, in Salt Lake County, Utah; and for a subset of tested patients, we performed manual chart review to examine symptoms and exposure risks. To determine risk factors for testing positive, we used logistic regression to calculate the odds of testing positive, adjusting for symptoms and prior exposure. Of the 20,088 individuals, 1,229 (6.1%) tested positive for SARS-CoV-2. We found that Non-White persons were more likely to test positive compared to non-Hispanic Whites (adjOR=1.1, 95% CI: 0.8, 1.6), and that this increased risk is more pronounced among Hispanic or Latino persons (adjOR=2.0, 95%CI: 1.3, 3.1). However, we did not find differences in the duration of symptoms nor type of symptom presentation between non-Hispanic White and non-White individuals. We found that risk of hospitalization increases with age (adjOR=6.9 95% CI: 2.1, 22.5 for age 60+ compared to 0-19), and additionally show that younger individuals (aged 0-19), were underrepresented both in overall rates of testing as well as rates of testing positive. We did not find major race/ethnic differences in hospitalization rates. In this analysis of predominantly non-hospitalized individuals tested for SARS-CoV-2, enabled by expansive testing capacity, we found disparities in both testing and SARS-CoV-2 infection status by race/ethnicity and by age. Further work on addressing racial and ethnic disparities, particularly among Hispanic/Latino communities (where SARS-CoV-2 may be spreading more rapidly due to increased exposure and comparatively reduced testing), will be needed to effectively combat COVID-19 in the US.

## Introduction

Since its emergence in humans in late 2019, coronavirus disease 2019 (COVID-19) (caused by severe acute respiratory syndrome coronavirus 2 (SARS-CoV-2) infection), has caused a global pandemic with over 3 million infections and 230,000 deaths. Currently, the United States (US) is the epicenter with approximately 30% of all global cases ^1^. Although the majority of case- and disease-course descriptions have originated outside of the US ^2-5^, emerging data suggests symptoms, risk from comorbidities, as well as age and racial patterns of COVID-19 may differ in the US compared to other highly-impacted countries such as China and Italy. The majority of US-based clinical or epidemiological descriptions of COVID-19 have been among hospitalized and ICU patients, due to limited testing capacity in much of the US ^6-11^. This bias in study population is potentially masking differences between hospitalized and non-hospitalized SARS-CoV-2 infections. Furthermore, due to the locations of most outbreaks, most US-based studies have focused on highly urban, densely populated areas with high levels of SARS-CoV-2 infection and relatively low testing capacity ^10-13^.

Widespread lack of SARS-CoV-2 testing capacity in the US has led to delays in case detection and isolation, significant under-reporting of cases (particularly those with mild and moderate symptoms), and gross underestimates of the true number of fatalities ^13-18^. In contrast, Utah is one of only two states (as well as Tennessee) that has maintained high per capita testing rates (testing 188 per 100,000 persons per day) as well as a stable proportion of positive tests (approximately 5.5% of all tests were positive since 12 March) (Figure 1)^19^. As such, testing in Utah has not been restricted to the most critical patients, and the majority of SARS-CoV-2 tests are administered in outpatient settings to community-dwelling, social distancing persons.

**Figure 1:**
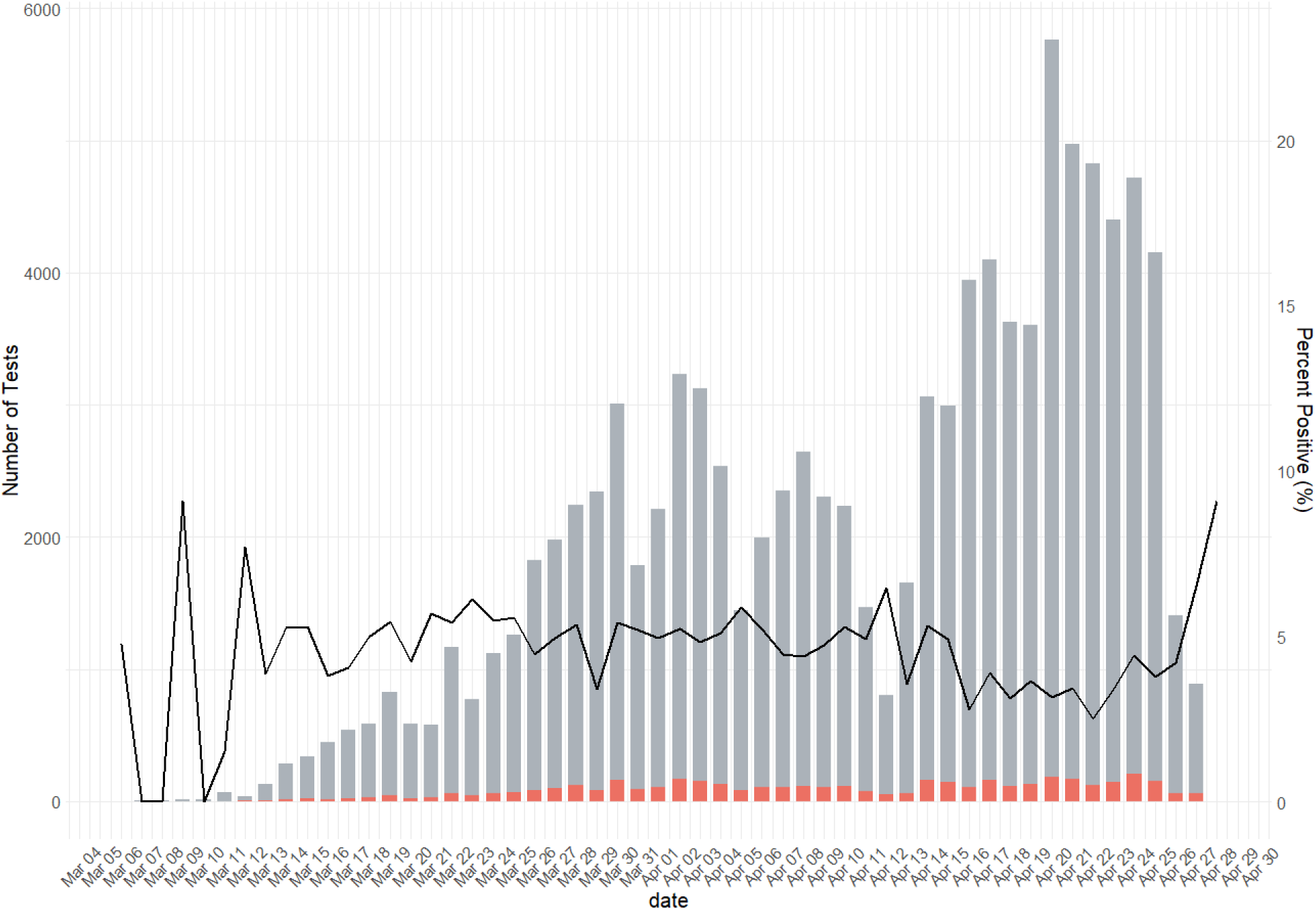
State-level SARS-CoV-2 testing data in Utah from 04 March to 28 April 2020. Grey bars represent the total number of tests performed each day in Utah and the red bars represent the total number of positive tests, based on the date they were ordered. The black line represents the percent of tests state wide that were positive each day.

The objective of this study is to compare demographic characteristics of positive and negative patients in a population for which testing is more readily available than previously published cohorts. In this article, we present the clinical, epidemiological, and demographic characteristics of all persons (hospitalized and community-dwelling) tested for SARS-CoV-2 infection at University of Utah Health (UHealth) from 10 March to 24 April 2020. UHealth primarily serves Salt Lake County, a diverse, medium-density, medium-sized metropolitan area of just over 1 million people. Due to the expanded testing capacity for SARS-CoV-2 in Utah, these data provide an alternative picture of the full spectrum of SARS-CoV-2 epidemiology relative to studies of hospitalized patients.

## Methods

All patients tested for SARS-CoV-2 in the UHealth system were eligible for our study. We developed an electronic, near real-time registry of all patients tested for SARS-CoV-2 at UHealth prior to 30 April 2020, for a total sample size of 20,088, representing approximately 20% of all tests conducted in the State of Utah during that time. The registry was built from the hospital operations dashboards, which contain basic demographics and test results for all patients tested for SARS-CoV-2. We linked this data with additional data using the Enterprise Data Warehouse (EDW) at UHealth. The EDW aggregates data from the health system’s disparate data collection systems in support of operations and research. We extracted demographics, International Classification of Diseases (ICD) billing codes, and vital signs, from the EDW to create the data set for this project. Demographic characteristics, including age, sex, and race/ethnicity were collected from all patients on intake for primary care or hospital visits, and via questionnaire for mobile testing centers. The department that ordered SARS-CoV-2 test was recorded, and data on comorbidities were collected via billing codes for all SARS-CoV-2 tested patients. While we were unable to determine if individuals were hospitalized as a result of their SARS-CoV-2 infection, we approximated this by determining if each person tested was hospitalized for any cause within 14 days of SARS-CoV-2 testing.

For each demographic variable, we calculated the odds ratios for testing SARS-COV-2 positive for a series of variables including race or ethnicity, age, and sex, adjusted for cough, fever, shortness of breath, and contact with a known SARS-COV-2 cases. Race/ethnicity was measured by comparing non-Hispanic White persons to non-White persons. Non-white persons included Hispanic/Latino, Asian, Black or African American, Native Hawaiian or other Pacific Islander, American Indian/Native Alaskan and other race individuals, as well as those with unknown race and those who chose not to disclose their race. We also compared Hispanic persons to non-Hispanic White persons, as they are the largest minority group in Salt Lake County and comprise 19% of the population.

We reviewed clinical text collected between 24 hours before and after SARS-CoV-2 testing for a subset of 2,043 persons tested prior to 31 March 2020 for additional manual chart review. Of the 20,088 total tests performed, 7,113 were performed prior to 31 March 2020. We selected a random subset, stratified by date, such that we included at least 20% of patients presenting on each day. This resulted in a sample size of 2043. Data on symptoms were extracted from the subset of manually reviewed charts. Symptoms were selected based on previous literature of SARS-CoV-2-positive patients ^20^. Epidemiologic risk factors collected from these charts included history of travel if mentioned, exposure to a SARS-CoV-2-positive person, healthcare worker status, and smoking status. Symptoms/exposure factors not mentioned in the medical chart notes were recorded as ‘not mentioned.’ In analysis we assumed “not mentioned” symptoms were absent. Percentage of present symptoms are seen below (Table 2).

In addition to collecting UHealth data, we extracted county-level and state-level data from the Utah COVID-19 dashboard for comparison ^19^.

This study was reviewed by the University of Utah Institutional Review Board (IRB) and determined to be exempt.

## Results

### Demographie Characteristics of Tested Individuals

Of the 20,088 patients tested for SARS-CoV-2 between 10 March 2020 and 24 April 2020, 1,229 (6.1%) tested positive for SARS-CoV-2. Testing occurred at 10 different types of facilities, including 18,399 (92%) in outpatient clinics, 279 (1%) at mobile testing sites, 958 (5%) in the emergency department (ED), 219 (1%) in an acute care facility, and 107 (0.5%) tested in the intensive care unit (ICU). The proportion of positive tests were similar between facility types (Table 1).

**Table 1:** Characteristics of SARS-CoV-2-TestedPatients (n=20088)

|  | **SARS-CoV-2**<br>**Positive**<br>**(N=1229)** | **SARS-CoV-2**<br>**Negative**<br>**(N=18859)** | **Adjusted<sup>1</sup>**<br>**Odds Ratio**<br>**(95% CI)** |
| --- | --- | --- | --- |
| **Age** |  |  |  |
| Median – (IQR) | 38.4 (27.6 - 51) | 39.2 (27.8 - 53.6) |  |
| 0-19 – no. (%) | 91 (7.4%) | 1745 (9.3%) | Reference group |
| 20-59 – no. (%) | 994 (81%) | 13956 (74%) | 1.6 (0.7, 3.6) |
| 60+ – no. (%) | 137 (11%) | 3154 (17%) | 1.4 (0.6, 3.4) |
| Missing – no. (%) | 7 (0.57%) | 4 (0.02%) | --- |
| **Sex** |  |  |  |
| Female – no. (%) | 545 (44%) | 10526 (56%) | Reference group |
| Male – no. (%) | 683 (56%) | 8323 (44%) | 1.4 (1.0, 1.9) |
| **Race/Ethnicity** |  |  |  |
| White non-Hispanic – no. (%) | 489 (40%) | 12662 (67%) | Reference group |
| Hispanic/Latino – no. (%) | 402 (33%) | 2402 (12%) | 2.0 (1.3, 3.1) |
| Unknown – no. (%) | 182 (15%) | 1739 (9.2%) | 0.8 (0.4, 1.6) |
| COMBINED<sup>2</sup> |  |  | 0.8 (0.4, 1.4) |
| Black/African American – no. (%) | 31 (2.5%) | 330 (1.8%) | --- |
| Asian – no. (%) | 30 (2.4%) | 416 (2.2%) | --- |
| American Indian and Alaska Native – no. (%) | 6 (0.49%) | 146 (0.77%) | --- |
| Native Hawaiian and Other Pacific Islander – no. (%) | 10 (0.8%) | 213 (1.1%) | --- |
| Other – no. (%) | 44 (3.6%) | 498 (2.6%) | --- |
| Refuse – no. (%) | 35 (2.9%) | 453 (2.4%) | --- |
| **BMI** |  |  | Not Calculated |
| Median – (IQR) | 30.1 (26 - 34.9) | 27.6 (23.2 - 32.9) | --- |
| Missing – no. (%) | 1161 (95%) | 17433 (92%) | --- |
| **Comorbidities** |  |  | Not Calculated |
| Any comorbidity | 134 (11%) | 3402 (18%) | --- |
| fluid/electrolyte disorders – no. (%) | 7 (0.57%) | 221 (1.2%) | --- |
| Renal Failure – no. (%) | 8 (0.65%) | 172 (0.91%) | --- |
| Alcohol Abuse – no. (%) | 5 (0.41%) | 207 (1.1%) | --- |
| Chronic blood loss anemia – no. (%) | 5 (0.41%) | 160 (0.85%) | --- |
| Chronic pulmonary disease – no. (%) | 10 (0.81%) | 220 (1.2%) | --- |
| Depression – no. (%) | 5 (0.41%) | 62 (0.33%) | --- |
| Hypertensive renal disease without renal failure – no. (%) | 14 (1.2%) | 398 (2.1%) | --- |
| Hypertension, uncomplicated – no. (%) | 111 (9.0%) | 2809 (15%) | --- |
| Obesity – no. (%) | 11 (0.90%) | 340 (1.8%) | --- |
| Other – no. (%) | 12 (0.98%) | 600 (3.2%) | --- |
| **Pulse Rate** |  |  | Not Calculated |
| Median – (IQR) | 87 (78 – 99) | 85 (75 - 96) | --- |
| Missing – no. (%) | 138 (11%) | 2068 (11%) | --- |
| **Respiratory Rate** |  |  | Not Calculated |
| Median – (IQR) | 18 (16 - 20) | 18 (16 - 20) | --- |
| Missing – no. (%) | 1108 (90%) | 16552 (88%) | --- |
| **SpO2** |  |  | Not Calculated |
| Median – (IQR) | 96 (94 – 97) | 96 (95 - 98) | --- |
| Missing – no. (%) | 131 (10.7%) | 2054 (11%) | --- |
| **Body Temperature (C)** |  |  | Not Calculated |
| Median – (IQR) | 37.1 (36.7 - 37.6) | 37 (36.6 - 37.3) | --- |
| Missing – no. (%) | 762 (62%) | 11118 (59%) | --- |
| **Facility that ordered SARS-CoV-2 test** |  |  | Not Calculated |
| Acute care – no. (%) | 7 (0.57%) | 212 (1.12%) | --- |
| ED – no. (%) | 55 (4.48%) | 903 (4.79%) | --- |
| External – no. (%) | 5 (0.41%) | 0 (0%) | --- |
| ICU – no. (%) | 6 (0.49%) | 101 (0.54%) | --- |
| Inpatient rehab – no. (%) | 0 (0%) | 10 (0.05%) | --- |
| LND/MNB – no. (%) | 0 (0%) | 34 (0.18%) | --- |
| Mobile – no. (%) | 230 (1.22%) | 230 (1.2%) | --- |
| Outpatient Clinics – no. (%) | 1095 (89.1%) | 17304 (92%) | --- |
| Telemedicine – no. (%) | 0 (0%) | 25 (0.13%) | --- |
| Other – no. (%) | 12 (0.98%) | 40 (0.21%) | --- |

<sup>1</sup>Adjusted for cough, fever, shortness of breath, and known contact with SARS-COV-2 case
<sup>2</sup>Due to lack of model convergence due to small cell counts, the following race/ethnicity variables were combined for odds ratio calculations: Black/African American, Asian, American Indian and Alaska Native, Native Hawaiian and Other Pacific Islander, Other, Refuse

**Table 2:** Symptoms in Subset of SARS-CoV-2-tested Patients (n=2043)

|  | SARS-CoV-2<br>Positive<br>(n=136) | SARS-CoV-2<br>Negative<br>(n=1907) |
| --- | --- | --- |
| **Symptoms** |  |  |
| Cough – no. (%) | 121 (89%) | 1697 (89%) |
| Fever – no. (%) | 103 (76%) | 1229 (64%) |
| Shortness of breath – no. (%) | 68 (50%) | 1239 (65%) |
| Myalgia – no. (%) | 57 (42%) | 572 (30%) |
| Headache – no. (%) | 50 (37%) | 462 (24%) |
| Lethargy – no. (%) | 43 (32%) | 496 (26%) |
| Nasal – no. (%) | 44 (32%) | 562 (29%) |
| sore throat – no. (%) | 41 (30%) | 592 (31%) |
| Diarrhea – no. (%) | 16 (12%) | 188 (9.9%) |
| nausea/vomit – no. (%) | 10 (7.4%) | 163 (8.6%) |
| Duration of symptoms – median (IQR) | 4.0 (2.3, 7.0) | 4.0 (2.0, 7.0) |
| **BEHAVIORS** |  |  |
| Prior exposure – no. (%) | 77 (57%) | 546 (29%) |
| Travel – no. (%) | 63 (46%) | 461 (24%) |
| Healthcare worker – no. (%) | 12 (8.8%) | 205 (11%) |
| Smoker – no. (%) |  |  |
| never – no. (%) | 78 (57%) | 869 (46%) |
| former – no. (%) | 5 (3.7%) | 164 (8.6%) |
| current – no. (%) | 4 (2.9%) | 186 (9.8%) |
| missing – no. (%) | 49 (36%) | 688 (36%) |

#### Race and Ethnicity

While the racial/ethnic distribution of the population tested at UHealth was broadly similar to the racial/ethnic distribution of Salt Lake County (Figure 2B), there were still statistical differences (p<0.001). Discrepancies were especially pronounced for persons identifying as Hispanic or Latino; Hispanic/Latino persons comprise 19% of the county population but only 10% of all tested at UHealth. Furthermore, we found that non-White persons were more likely to test positive compared to non-Hispanic White/Caucasians (adjOR=1.1, 95% CI: 0.8, 1.6), and Hispanic/Latino persons were more likely to test positive compared to non-Hispanic White/Caucasian persons (adjOR=2.0, 95% CI: 1.3, 3.1; Table 1, Figure 2). Likewise, when comparing the location where individuals presented for testing (e.g., ED, outpatient clinic, mobile testing site), we found variation by race/ethnicity (p<0.001), but Hispanic/Latino persons were only slightly more likely to be tested in the ED compared to non-Hispanic White/Caucasian (6.3% versus 4.9%). We did not find a difference in reported symptoms between Hispanic/Latino and non-Hispanic White/Caucasian persons.

**Figure 2:**
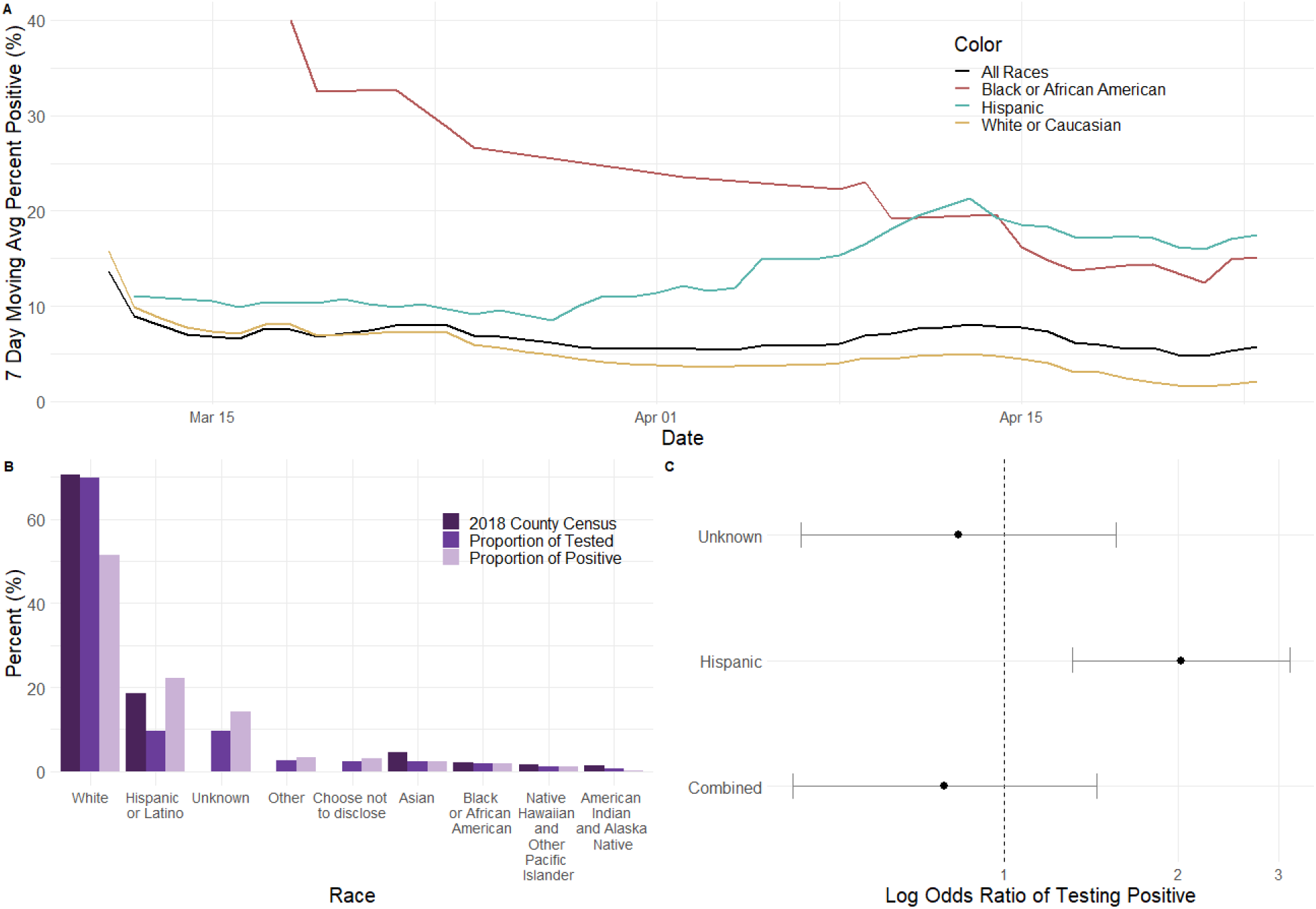
Epidemiological charactenstics of persons who were tested for SARS-CoV-2 at UHealth by race/ethnicity. A) Shows 7-day moving average of the percent oftests that were positive by race/ethnicity, each day over the course of the study, for select race and ethnicities. Red, teal, and yellow represent the percent positive for Black or African American, Hispanic, and White or Caucasian persons while black represents the 7-day moving average across all races. B) Shows the percent of persons by each race from the 2018 census estimates for Salt Lake County^34^, the proportion of persons tested by race/ethnicity, as well as the proportion of persons who tested positive by race/ethnicity C) The log-odds of testing positive by race, compared to White/Caucasian. Adjusted for cough, fever, shortness of breath, and contact with known SARS-COV-2 case. Due to lack of model convergence due to small cell counts, the following race/ethnicity variables were combined for odds ratio calculations: Black/African American, Asian, American Indian and Alaska Native, Native Hawaiian and Other Pacific Islander, Other, Refuse

#### Age Structure

The population tested at UHealth is not similar to the age structure of Salt Lake County (Figure 3). However, the age distribution of those tested at UHealth is consistent between individuals testing negative versus positive. Younger individuals (aged 0-19), were underrepresented in overall rates of testing and rates of testing positive (Table 1, Figure 3). Without adjusting for the underlying age distribution of the county population, we found that persons aged 20-59 had the highest odds of testing positive compared to those 0-19 years old (adjOR=1.6, 95% CI: 0.7, 3.6, Table 1).

**Figure 3:**
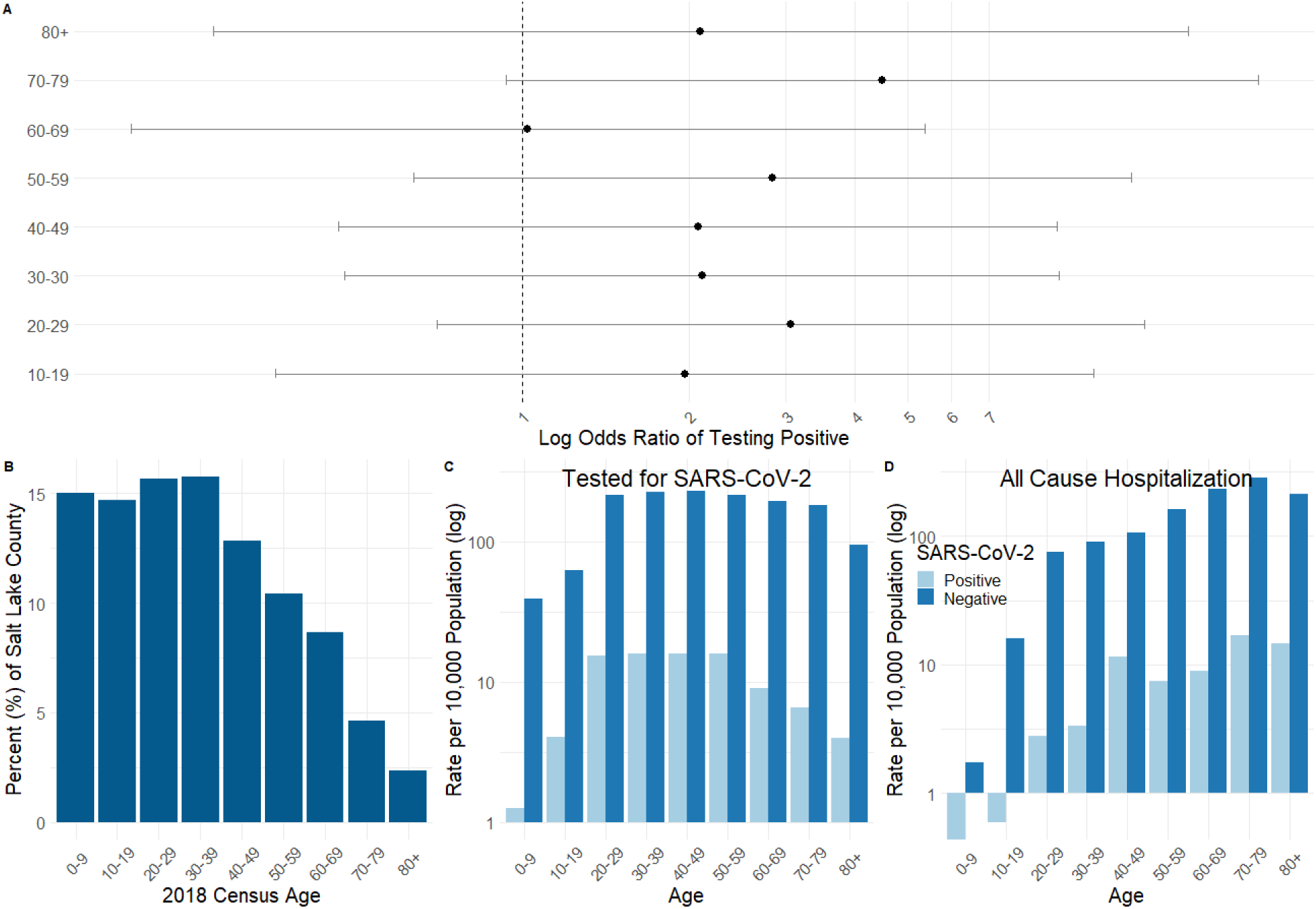
Epidemiological characteristics of persons who were tested for SARS-CoV-2 at UHealth by age. A) The log-odds of testing positive by age, compared to 0-9 year olds. Adjusted for cough, fever, shortness of breath, and contact with known SARS-COV-2 case. B) Shows the percent of persons in each age group from the 2018 census estimates for Salt Lake County^34^. C) The rate of testing, per 10,000 population, by age for persons testing (light blue) SARS-CoV-2 positive and (dark blue) SARS-CoV-2 negative. D) The rate of testing, per 10,000population, by age all cause hospitalizations within 14 days of testing for (light blue) SARS-CoV-2 positive and (dark blue) SARS-CoV-2 negative.

#### Sex

Persons who tested positive for SARS-CoV-2 were more likely to be male. Among individuals who tested positive for SARS-CoV-2, over 56% were male (44% were female), while 44% of individuals who tested negative were male (56% of female). Excluding individuals for which gender was unknown or non-binary (11 people), males had 1.4 times the odds of testing SARS-COV-2 positive compared to females (95% CI: 1.0, 1.9).

### Clinical Characteristics

Among manually extracted charts, 136 (6.7%) individuals tested positive for SARS-CoV-2 between 10 March 2020 and 31 March 2020 (Table 2). This was similar to the proportion of individuals testing positive for SARS-CoV-2 among all those tested 1229 (6.5%).

#### Clinical symptoms

Persons testing SARS-COV-2-positive and SARS-COV-2-negative had similar vital signs and rates of cough (89%), but varied in presentation of other symptoms. Symptom profiles were different in persons who tested SARS-COV-2 positive compared to those who tested SARS-COV-2 negative (Table 2). The median duration between symptom onset and presentation for testing was 4 days (IQR=2.1, 7.0) for both SARS-COV-2 positive and SARS-COV-2 negative persons (Table 2).

#### Hospitalization rates

Using all-cause hospitalization occurrence within 14 days after testing, we found that 5.1% of persons tested positive were hospitalized, and 5.1% of persons testing negative were hospitalized. Rates of hospitalization were similar across racial ethnic groups, with 6% of people identifying as Black/African American, 5% of people identifying as non-Hispanic White/Caucasian, and 5% of people identifying as Hispanic/Latino being hospitalized within 14 days of SARS-CoV-2 testing. Those identifying as American Indian and Alaskan Native had higher rates of 14-day hospitalization (9%), but the sample size was limited.

We found that hospitalization differed by age, and older adults had the highest odds of all-cause hospitalization (adjOR=6.9, 95% CI: 2.1, 22.5 for adults 60+ compared to 0-19-year olds; Table 1, Figure 3). Additionally, we found all-cause hospitalization rates increased with age in both SARS-COV-2-positive and SARS-COV-2-negative persons (Figure 3d).

#### Epidemiological Risk Factors

Persons testing SARS-CoV-2 positive were more likely to have known exposure to another SARS-CoV-2 person or a history of travel (Table 2). Among SARS-COV-2-positive persons, 57% had been in contact with a confirmed SARS-CoV-2 case and 46% had travelled before their own SARS-CoV-2 testing, compared to 29% and 24% in SARS-COV-2-negative persons, respectively (Table 2). 9% of SARS-COV-2-positive persons and 11% of SARS-COV-2- negative persons were healthcare workers (Table 2).

## Conclusions

The high-level of SARS-CoV-2 testing availability in Utah has allowed for a higher proportion of the outpatient, community dwelling (and usually social distancing) population to be tested. This expands on previous studies that were limited to more severe or critically-ill patients, as a result of limited test availability. These data provide insight into the epidemiological and clinical characteristics of COVID-19 over a much broader range of disease severity, as well as highlight disparities in COVID-19 response. While the data presented in this study represents only 20% of the total number of people tested in Utah, based on publicly available state-level data, our findings are largely representative of the overall state-level outbreak.

While the racial/ethnic makeup of the population tested for SARS-CoV-2 is broadly similar to overall county demographics (Figure 2), important discrepancies remain. We found that persons who identified as non-White had elevated odds of testing positive compared to non-Hispanic White/Caucasians (adjOR=1.1, 95% CI: 0.8, 1.6). The discrepancy was especially pronounced for persons identifying as Hispanic or Latino, who had 2.0 times the odds (95% CI: 1.3, 3.1) of testing SARS-CoV-2 positive compared to those who identified as non-Hispanic White (Figure 2). Though our study is limited to those within a single health system, our results are representative to those of the county and the state as a whole. For example, Hispanics make up 10% of our tested population, and 22% of those testing positive, which is broadly consistent with state-level data in which Hispanics make up 14.2% of the population and 36.6% of those testing positive ^19^.

There are likely several factors that contribute to the disparities in test-positive rate ^21^. Hispanic/Latino persons are disproportionally employed as essential workers, which increases exposure risk. While Hispanics make up 17.6% of the US workforce, they comprise 25% of service occupations ^22^. This increased employment exposure is likely compounded by the lack of paid sick leave, as Hispanic workers have lower rates of access to sick leave than do White workers ^23^. Workers without paid sick leave are likely to continue working even while sick, leading to increased exposure to co-workers and family-members. Another compounding factor is the decreased ability for public health personnel to conduct contact tracing within minority communities due to historical and current political instances leading to lack of trust between minority communities and government bodies ^24-29^. It is also the case that Hispanic workers may be less able to stop working and stay home due to low incomes. Hispanic households in Salt Lake County make $56,498/year as compared to $82,026 among non-Hispanic White households ^30^. Lower incomes also translate into more crowded housing situations, which facilitate virus transmission. In the US, 10% of Hispanics live in overcrowded conditions ^31^ and that figure rises to 33% among older Hispanics in the urban US^32^.

Persons identifying as non-Hispanic White/Caucasian and Hispanic/Latino were tested at each facility at approximately equal rates – approximately 5% were tested in the ED, and 90% in outpatient clinics. Likewise, using symptom presentation or hypoxia (SpO2 < 90%) as a proxy for severity, we find no difference between persons identifying as non-Hispanic White/Caucasian and as Hispanic/Latino. The duration of time from symptom onset to seeking testing was also similar across races. This suggests racial disparities in SARS-COV-2 burden are not simply due to Hispanic/Latino persons delaying care until symptoms are critical and presenting to the ED, nor is it as straight forward as preferentially presenting to mobile testing centers. Identifying the underlying cause of these health disparities is a critical step towards containing SARS-CoV-2.

Unlike previous studies ^6-11^ that focus only on severe or critical patients, we are also able to identify differential patterns of testing by age. Promisingly, we find most adults (aged 20-79) in our study have high testing rates (over 100 tests per 10,000 population), which indicates that we are capturing large numbers of mild and moderate illness. On the other hand, we also show that testing rates remain lower in children (under 20 years old), and of those under 20 years old who are tested, rates of SARS-CoV-2 positivity are disproportionately lower. This supports previous studies which highlight risk of asymptomatic SARS-CoV-2 in younger individuals, as inclusion in our study was predicated by having at least one symptom (cough, fever, or shortness of breath)^20^.

In addition to differences due to race, we also identified differences due to age. We found the highest hospitalization rates were among middle-aged SARS-COV-2-positive patients (Figure 3). This is broadly consistent with Myers et al. ^6^ who found hospitalized and ICU admitted patients in California tended to be middle-aged. Like similar studies^8^, we found hospitalization rates were smallest among younger age individuals (aged 0-19). Indeed, we found persons under 20 years old who tested positive for SARS-CoV-2 were far less likely to be hospitalized than their peers who were tested for SARS-CoV-2 but tested negative. Compared to other studies, we found age-adjusted lower hospitalization rates than previously reported among those who test positive.

Given that Utah has the youngest population in the country ^33^, a disease that is more likely to be asymptomatic in younger people is especially salient for the overall state burden and the ability to control the outbreak as social distancing measures are loosened. Given our finding that younger individuals are underrepresented in testing due in part to testing criteria, state-level strategies should be re-examined to consider strategies to target testing of younger age groups based on geospatial and epidemiological risk factors, regardless of symptomatology.

A benefit of expanded testing locations in Salt Lake County was that we were able to assess the demographic characteristics of testing locations. We found that those tested in outpatient facilities tended to be younger and that the proportion of persons tested in the ED increased with age. We found no difference in race/ethnicity by testing location. If testing were to be expanded to lift symptom requirements in an effort to identify more mild and asymptomatic infections, our study highlights the benefit of outpatient and mobile facilities to reach underrepresented demographics.

This study offers an important contribution to existing COVID-19 literature, which has been primarily focused on hospitalized patients. In contrast to previous work, over 90% of people included in our study were tested in outpatient or community settings. However, our data is limited to persons who met testing criteria; very few asymptomatic infections are captured in this study. Further work understanding the demographic and epidemiological characteristics of asymptomatic infections remains vital to understanding and controlling SARS-CoV-2. By highlighting critical gaps in testing, particularly among Hispanic/Latino communities, where SARS-CoV-2 may be spreading more rapidly due to increased exposure and comparatively reduced testing, this study takes a critical first step towards reversing these health disparities. Our findings, based largely on those who received testing as outpatients, highlights the potential gaps in control of SARS-CoV-2 infection related to age, race, and ethnicity.

## Data Availability

Data contain identifiable information and therefore will not become public

